# Assessment of the variant prioritisation strategy for genomic newborn screening in the Generation Study

**DOI:** 10.1101/2025.03.12.25323745

**Authors:** Joanna Kaplanis, Dasha Deen, Prasanth Sivakumar, Mafalda de Almeida Gomes, Arina Puzriakova, Ivone Leong, Kevin Savage, Gabriel Aldam, James Skelton, Paul Quinn, Andrew Parrish, Emma Baple, Meekai To, Katrina Stone, David Bick, Amanda Pichini, Alice Tuff-Lacey, Richard Scott, Ellen Thomas, Dalia Kasperaviciute

**Affiliations:** Genomics England, London, UK; Genomics Laboratory, Royal Devon and Exeter NHS Foundation Trust, Exeter, UK; South West Genomic Medicine Service, England, UK; Department of Clinical and Biomedical Sciences, University of Exeter Medical School, Exeter, UK; Peninsula Clinical Genetics Service, Exeter, UK; Harris Birthright Centre, King’s College Hospital, London, UK; Genetics and Genomic Medicine Department, UCL Great Ormond Street Institute of Child Health, London, UK; Clinical Genetics Department, Great Ormond Street Hospital for Children, London, UK

## Abstract

**Purpose:** Genomic sequencing offers the opportunity to screen for hundreds of rare genetic conditions with a single test. To minimise potential negative impact on families and clinical services, it is crucial to reduce false positive results while prioritising clinical utility. Here we present an automated variant prioritisation approach for genomic newborn screening and the assessment of its validity across the conditions included in the Generation Study, a research study investigating genomic sequencing in 100,000 newborns in England. Prioritised variants will subsequently undergo manual review by a registered clinical scientist, and a specialist clinician, before being reported back to parents.

**Methods:** The automated variant prioritisation approach was evaluated for its ability to predict the presence or absence of relevant genetic variants, subsequently informing gene and variant-specific inclusion in the study. Specificity was estimated using a cohort of 34,410 samples not enriched for rare diseases, while sensitivity was assessed with 546 samples from participants with diagnostic variants in genes relevant to newborn screening. Coverage and CNV callability metrics were evaluated as proxies for variant detection.

**Results:** Assessing validity of automated variant prioritisation on a gene level led to changes in rules used in variant prioritisation and conditions included. We estimated that 3-5% of samples will have prioritised variants that require manual review and that <1% of samples will be taken forward for orthogonal confirmatory testing and genetic confirmation. We prioritised variants in ∼80% of samples with diagnostic variants across genes included in the Generation Study.

**Conclusion:** Gene-specific assessment of variant prioritisation is crucial to establish analytical validity prior to inclusion in genomic newborn screening.

## Introduction

Genomic sequencing offers the opportunity to screen for hundreds of rare conditions with a single test, enabling earlier treatment^1,2^. Multiple research initiatives worldwide are exploring the potential benefits and challenges of genomic newborn screening (gNBS)^3–15^. Most studies use automated pipelines to prioritise relevant variants, followed by manual review, though the number of variants reviewed manually varies. The Generation Study aims to recruit 100,000 newborn babies to evaluate the utility and feasibility of newborn screening using genome sequencing (GS), understand how the genomic data can be used for research and explore the potential risks and benefits of storing and reusing a baby’s genome over their lifetime. Led by Genomics England, in partnership with the National Health Service (NHS), the Generation study will be assessed for its utility alongside the current UK Newborn Blood spot test that screens for nine conditions. The study includes 209 severe, early-onset conditions caused by genetic variants in 463 genes where early, equitably accessible interventions could improve outcomes.

Genomic newborn screening poses challenges due to limited understanding about the pathogenicity, expressivity and penetrance of genetic variation, particularly without clinical phenotypes. In this context, it is paramount to minimise the number of false positive results reported to participants to reduce potential harmful impact on families and overburdening clinical services. Moreover, assessing the clinical relevance of genetic variants across many conditions at scale can be time-consuming and costly, necessitating careful resource planning, particularly within a publicly funded national health system. To address this, we developed an automated approach that prioritises variants, followed by manual review by a clinical scientist, a registered healthcare professional with expertise interpreting genomic variation. This approach aims to maximise the positive predictive value of results reported to the participants while minimising the number of variants requiring review. We also aim to reduce the prioritisation of variants not relevant to newborn screening, such as those associated with adult-onset conditions or heterozygous states for recessive conditions. The pipeline was evaluated for its ability to predict the presence or absence of relevant genetic variants and inform gene inclusion in the study (Figure 1). Specificity was estimated using 34,410 participants not enriched for rare diseases and sensitivity assessed using diagnostic variants from the 100,000 Genomes Project^16^ and NHS Genomic Medicine Service (GMS)^17^. Coverage and CNV callability were also assessed as proxies for variant detection per gene. Here, we describe the development and optimisation of this pipeline, including the challenges faced and solutions implemented.

**Figure 1:**
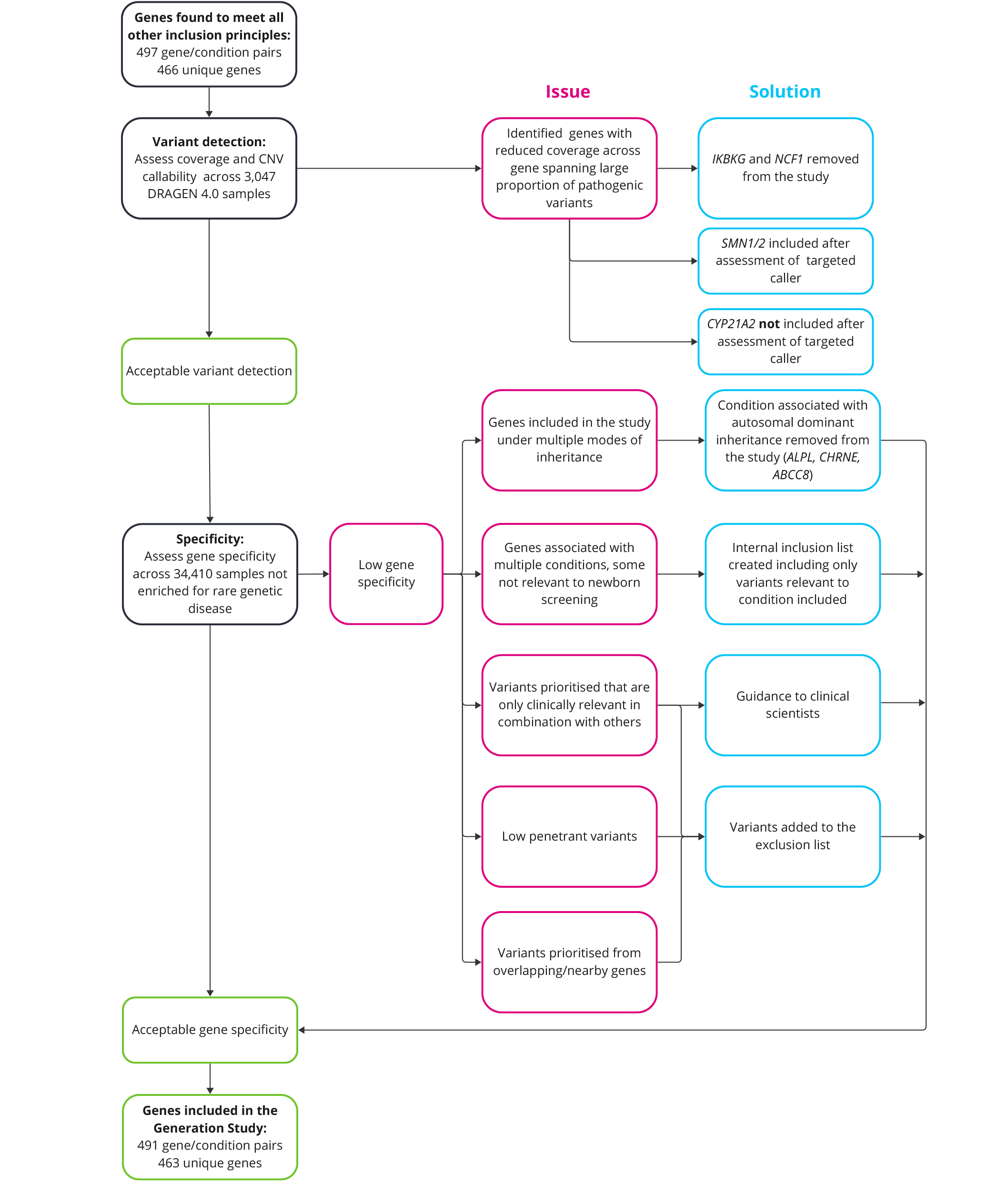
Overview of analytical validity evaluation for genes assessed for inclusion in the Generation Study.

## Methods

### Alignment and variant calling

The Newborn Screening Pipeline reports using genome reference GRCh38. DRAGEN v4.0.5 is used for alignment and variant calling, including small variants and copy number variants (CNVs). Variants are prioritised for chromosomes 1-22 and X in relevant genes. Copy number variants (CNVs) (>2kb) were detected using the DRAGEN CNV workflow with self-normalisation and the Shifting Level Models (SLM) segmentation mode are prioritised through the predicted loss of function (pLoF) algorithm. High-quality CNVs >10 kb are defined as those with filter status PASS. CNVs between 2-10 kb are identified by combining DRAGEN CNV workflow and DRAGEN SV caller results. CNVs detected by both with a minimum reciprocal overlap of 50% are considered high quality and set to PASS. Mitochondrial genes are not assessed, as they are not included in the screened conditions. Variant annotation is done using Cellbase (https://github.com/opencb/cellbase). Annotation of multi-nucleotide variants (MNVs) is supported when phase information is available (read-based phasing).

### Gene-condition pairs in the Generation Study

Initially, 922 gene-condition pairs were assessed against the four principles developed for Generation Study selection (Supplemental Table 1, https://www.genomicsengland.co.uk/initiatives/newborns/choosing-conditions). Reliable variant detection is fundamental to Principle A; that there is strong evidence the genetic variant causes the condition and can be reliably detected. Of these, 497 gene-condition pairs (466 unique genes) met all other principles and were brought forward to assess reliable variant detection (Supplemental Table 2).

### Cohorts used for analyses

Variant prioritisation was optimised and specificity was initially assessed using data from 34,410 participants (primarily adults), including 28,484 participants recruited and sequenced by Genomics England as part of the COVID-19 Genomics Study and 5,925 participants from the cancer arm of the 100,000 Genomes Project^18,19^. These were included in the aggregated dataset v5 data release (aggCOVIDV5; https://re-docs.genomicsengland.co.uk/covid5/). This included high-coverage short-read GS sequencing data aligned to the NCBI GRCh38 reference genome (mean autosomal read depth: ∼43x). Alignment and variant calling performed with DRAGEN v3.2. Selected to reflect the ancestry composition of the UK (Supplemental Table 3) and not expected to be enriched for rare diseases, this cohort will be referred to as the control-like cohort. A subset of 5,855 samples with mild COVID-19 or cancer from the control-like cohort were realigned using DRAGEN v4.0.4 to best represent the approach used in the Generation Study; this realigned subset is referred to as the control-like subset. This cohort was used to generate internal allele frequencies used in variant prioritisation.

Specificity validation was performed on a replication cohort of 1,362 participants with cancer, excluding haematological cancers, using germline samples from the NHS Genomics Medicine Service that consented to inclusion in the National Genomic Research Library (NGRL) ((*The National Genomic Research Library v5.1, Genomics England*. https://doi.org/10.6084/m9.figshare.4530893/7.2020). Alignment and variant calling was performed with DRAGEN v3.2.

Sensitivity was assessed using 546 participants with diagnostic small variants in genes included in the Generation Study. This comprised of 375 participants from the rare disease arm of the 100,000 Genomes

Project and 171 NHS Genomic Medicine Service participants that consented to inclusion in the NGRL. These were selected on the following criteria:

∘ The small variant was reported and case marked as solved
∘ Variants annotated as pathogenic or likely pathogenic
∘ Variants were found to fully explain phenotypes
∘ The mode of inheritance aligned with that included in the Generation Study
∘ Participants processed with genome assembly GRCh38

Participants age at recruitment ranged from 0-76 years and their phenotypes were not reviewed to confirm alignment with those included in the Generation Study. Samples were realigned with DRAGEN v4.0.5.

### Coverage and CNV callability analysis

Coverage was assessed in 3,047 female samples from the control-like cohort realigned with DRAGEN v4.0.4. Coverage metrics (mean, median and proportion >15X) were calculated in relevant transcripts (MANE Clinical/Select v1, otherwise Ensembl v107canonical) per transcript and per exon where Mapping Quality >10, Base Quality > 30 and soft clipped reads removed (Supplemental Table 4).

CNV callability was assessed by the proportion of the gene that falls into regions of increased sequence homology which are excluded by the DRAGEN v4 CNV caller. Genes that had compromised coverage CNV callability across the gene were removed from the Newborn gene list (*IKBKG* and *NCF1*) (Supplemental Figure 1). Genes were only excluded only if poor coverage affected the entire gene (median and mean coverage <30X and 95% of the gene with coverage <15X) or if CNV callability was compromised in genes where most known pathogenic variants are CNVs. Genes with poor coverage statistics but with targeted callers available were assessed separately (*SMN1, SMN2* and *CYP21A2*).

### Variant prioritisation for Newborn Screening

Variants are prioritised if previously classified as pathogenic/likely pathogenic for included conditions or if predicted to cause protein loss-of-function, when relevant for the screened condition (Figure 2). Data sources include ClinVar^20^, a curated dataset of clinical variants from QIAGEN Clinical Insights^20^, reported variants that have come through the NHS Genomic Medicine Service and 100,000 Genomes Project which are accessed through the Clinical Variant Ark (CVA) (https://ip-cva-help.genomicsengland.co.uk/) and internal inclusion lists compiled by Genomics England clinical and curation teams, with the input of external specialists (Supplemental Table 5). The rules described correspond to the first iteration of the pipeline, however these will evolve during the study. Resources and variant lists will continue to be updated.

**Figure 2:**
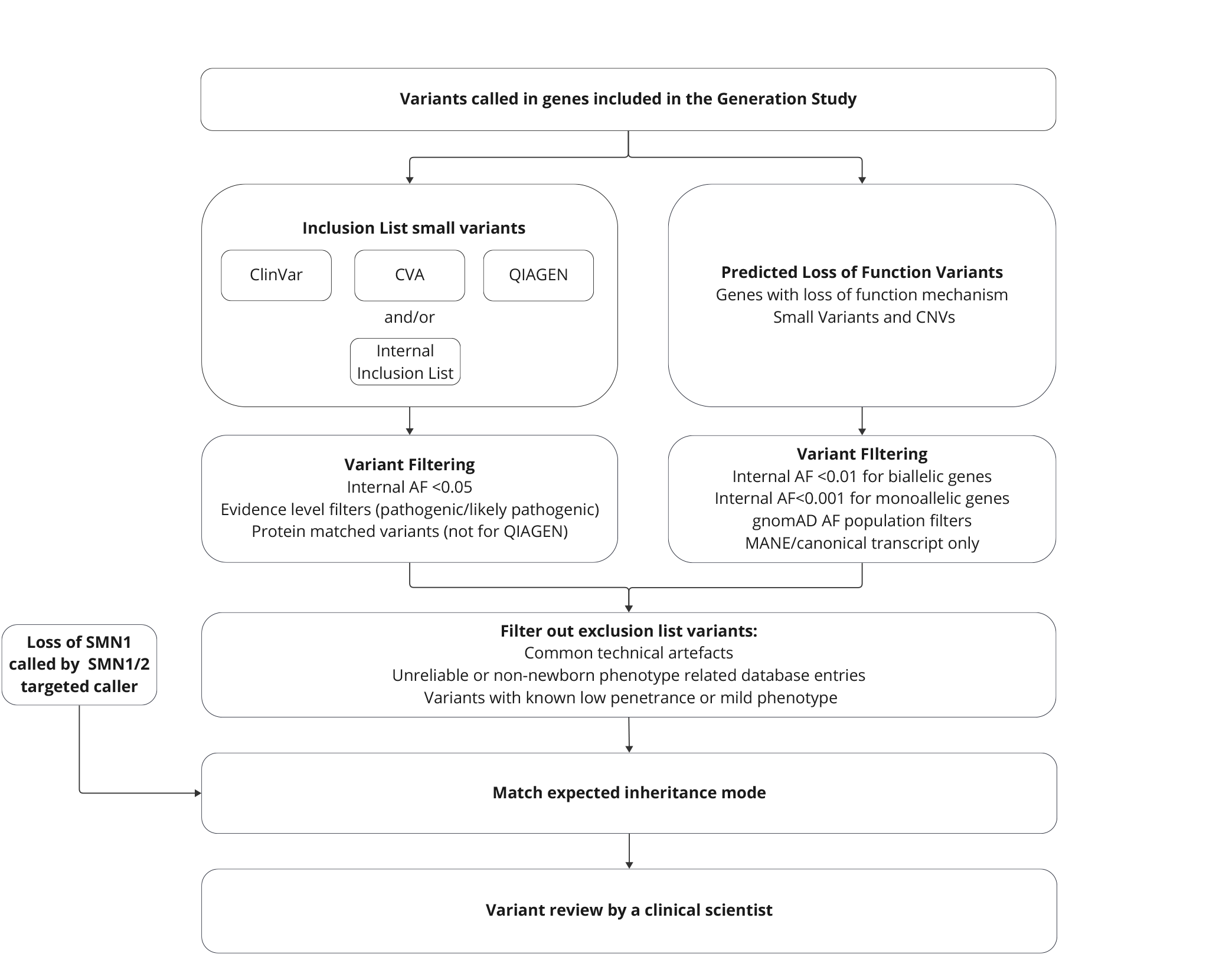
Overview of variant prioritisation approach in the Generation Study. Internal AF refers to allele frequencies derived from the control-like subset. Allele frequencies for gnomAD are detailed in Supplemental Table 8.

This section describes rules for the automated variant prioritisation, not guidelines for what is considered reportable. All prioritised variants in the Generation Study will be manually reviewed and only variants deemed clinically significant will be reported back to the specialist clinician and confirmatory testing and care pathways will be initiated when appropriate.

The following rules apply to all variants.

- Variants assigned PASS status for the DRAGEN quality threshold
- Variants match the expected inheritance mode of the condition included in the study. Read-based phasing allows for phasing of potential compound heterozygous variants within ∼150bp, however unphased variants that are potentially compound heterozygous will be prioritised.
- Variants will not be prioritised if they are on the exclusion list (Supplemental Table 6). This is a list maintained and curated by Genomics England which include common sequencing artefacts, low penetrant variants and variants associated with conditions or phenotypes not included in the study.

The further rules for filtering variants are specific to each of the potential sources of prioritisation and are described in the sections below.

#### Variants previously reported as pathogenic

- Only variants located within 50bp of a gene in the panel are considered for prioritisation.
- Variants present in the April 2023 ClinVar accession were prioritised under the following criteria:
  ∘ The variant or a variant with the same predicted protein change has at least one pathogenic/likely pathogenic classification (protein matching is not supported for MNVs).
  ∘ The variant has no more than one benign/likely benign classification
  ∘ Internal allele frequency < 0.05. This was applied to reduce potential submission errors or variant calling artefacts. For MNVs, the minimum allele frequency of the decomposed variants <0.05.
- Clinical Variant Ark (CVA) is Genomics England’s database of small variants with classifications and interpretation outcomes from the rare disease participants from the 100,000 Genomes Project and NHS GMS. These variants will be prioritised under the following criteria:
  ∘ The variant or a variant with the same predicted protein change has at least one pathogenic/likely pathogenic classification
  ∘ Internal allele frequency < 0.05.
- QIAGEN maintains a list of variants classified according to their own interpretations using the ACMG criteria. Data was analysed with QIAGEN Clinical Insight Interpret version 9.3.2 (https://digitalinsights.qiagen.com/). These variants will be prioritised under the following criteria:
  ∘ Internal allele frequency < 0.05.
  ∘ The variant is classified as pathogenic/likely pathogenic (protein matching will not be used for QIAGEN)
- Internal inclusion lists were compiled by Genomics England clinical and curation teams, with the input of external specialists. There are 12 genes that have an internal inclusion list and where other data sources will not be used (Supplemental Table 5,7). Additionally, for some genes, variants are included in the internal inclusion list alongside other data sources. Variants were required to have PubmedIDs as the evidence source to be included.

#### Prioritisation of predicted loss of function (pLoF) variants

Single nucleotide variants (SNVs) or small indels need to meet the following criteria:

∘ Annotated with high impact consequence type in MANE Select, MANE Clinical or predefined transcript where MANE v1.0 transcripts are unavailable. Predefined transcripts were chosen based on several criteria: the Ensembl Canonical tag and GENCODE Basic tags were used to identify suitable transcripts; the transcript with the longest coding sequence was prioritised to reduce the risk of missing pLoF variants; and published literature was reviewed to ensure alignment with phenotypes included in the study.
∘ Allele frequencies in all population datasets are below thresholds as specified in Supplementary Table 8. For MNVs, the minimum allele frequency of the decomposed variants is below all thresholds
∘ In a gene where LoF is determined to be the mode of pathogenicity for the condition included.

CNVs need to satisfy the following criteria to be prioritised:

1. The CNV +/-2kb must overlap the coding region of the gene if the gene with LoF pathogenicity is coding, or for non-coding genes, must overlap the transcript.
2. For CNV gains, both breakpoints must occur within the same transcript.
3. The CNV must be rare in the internal reference frequency data, using an 80% reciprocal overlap frequency. Frequency thresholds: CNV losses in dominant genes < 0.001, recessive genes < 0.005, CNV gains in dominant genes < 0.002, recessive genes < 0.01.
4. The CNV aligns with condition-associated mode of inheritance for LoF mode of pathogenicity.

#### SMN1/2

Details regarding the SMN Caller can be found in the DRAGEN v4.0 user guide (https://support-docs.illumina.com/SW/DRAGEN_v40/Content/SW/DRAGEN/SMNCaller.htm) and is derived from a method described in Chen X, Sanchis-Juan A, French CE, et al., 2020^21^. The case will be prioritised if inferred copy number of intact *SMN1* is zero.

### Specificity analyses

Specificity was calculated across all genes and per gene by running the control-like cohort aligned on DRAGEN v3.2 through the Newborns variant prioritisation pipeline excluding targeted callers (*SMN1/2, CYP21A2*). Specificity was subsequently estimated on the control-like subset realigned to DRAGEN v4 and the replication cohort (DRAGEN v3.2).

To assess the impact of ClinVar classifications filtering on specificity, we ran the pipeline on the control-like under different filters for variants with conflicting interpretation of pathogenicity (CIP):

- No prioritisation of CIP variants
- Prioritisation of CIP variants with ≥1 pathogenic/likely pathogenic and 0 benign/likely benign classifications
- Prioritisation of CIP variants with ≥1 pathogenic/likely pathogenic and ≤1 benign/likely benign classifications
- Prioritisation of CIP variants with ≤1 benign/likely benign classifications
- Prioritisation of all CIP variants

These scenarios were also applied to participants with diagnostic variants in study genes to assess sensitivity impact.

Potentially compound heterozygous variants were assessed using the gnomAD v3.2 co-occurrence tool to determine haplotype predictions. Variants were lifted over to GRCh37 with pyliftover and queried in the gnomAD browser^22,23^.

To evaluate if cancer susceptibility genes had more prioritised variants than expected in the replication cohort, we compared prioritised variants in newborn genes also appearing on NHS GMS cancer germline panels to those in other newborn genes. Cancer susceptibility panels included:

- Childhood solid tumours: https://panelapp.genomicsengland.co.uk/panels/243/
- Adult solid tumours cancer susceptibility: https://panelapp.genomicsengland.co.uk/panels/245/
- Haematological malignancies cancer susceptibility: https://panelapp.genomicsengland.co.uk/panels/59/

To estimate the number of reportable variants from the specificity analyses, prioritised variants were manually curated by a scientific curator, who reviewed literature to determine reportability, serving as a proxy for clinical scientist manual review in the study.

## Results

Modelling variant prioritisation on 34,410 individuals elucidates challenges in prioritising non-reportable variants

We applied our automated variant prioritisation approach on 34,410 participants from the control-like cohort, not expected to be enriched for rare disease (Figure 2). This was performed across 493 gene-condition pairs (462 unique genes) to assess their inclusion in the Generation Study. These genes met other inclusion principles, and variant detection was sufficient as judged by coverage and CNV callability assessment. Two genes, *IKBKG* and *NCF1* were excluded due to compromised coverage and CNV callability (see methods, Supplemental Table 4, Supplemental Figure 1). Specificity was calculated both per gene and across all genes (Table 1, Supplemental Figure 3, Supplemental Table 9). This analysis highlighted the following scenarios where many prioritised variants were not clinically relevant for newborn screening and are described below.

**Table 1:** Specificity of variant prioritisation across different cohorts not expected to be enriched for rare disease. Cohort details are described in the methods.

| Data source (DRAGEN version) | Data description | Sample Size | Prioritisation Source | Unique variants count | Samples with prioritised variant | Specificity (%) | 95% Confidence interval |
| --- | --- | --- | --- | --- | --- | --- | --- |
| Control-like cohort (DRAGEN v3.2) | Participants recruited for COVID-19 and from cancer arm of 100,000 Genomes Project | 34,410 | ClinVar | 349 | 811 | 97.64 | [96.4, 97.80] |
|  |  |  | Loss of Function | 348 | 459 | 98.67 | [98.54, 98.78] |
|  |  |  | QIAGEN | 303 | 600 | 98.26 | [98.11, 98.39] |
|  |  |  | CVA | 60 | 291 | 99.15 | [99.05, 99.25] |
|  |  |  | Internal Inclusion List | 7 | 13 | 99.96 | [99.94, 99.98] |
|  |  |  | All | 749 | 1289 | 96.25 | [96.05, 96.45] |
| Control-like subset (DRAGEN v4.0) | Participants recruited with mild COVID-19 and participants from cancer arm of 100,000 Genomes Project | 5,855 | All | 141 | 161 | 97.25 | [96.80, 97.64] |
| Replication cohort (DRAGEN v3.2) | Participants recruited from the NHS Cancer GMS | 1,362 | All | 80 | 62 | 95.45 | [94.21, 96.43] |

### Genes associated with multiple conditions or multiple mechanisms of disease

Genes associated with multiple conditions or disease mechanisms can lead to prioritisation of variants irrelevant for newborn screening. To address this, internal variant inclusion lists were compiled by Genomics England clinical and curation teams, with the input of external specialists. These have been created for 12 genes where databases contain many pathogenic/likely pathogenic variants for conditions or phenotypes not included in the study (Supplemental Table 5, 7). While specificity was already high for some genes, *CFTR* specificity notably increased from 97.81% to 99.97% after applying the internal inclusion list.

### Low penetrance variants

We implemented several strategies to reduce prioritisation of variants with reduced penetrance that are unsuitable for newborn screening. We modelled different ClinVar filtering criteria on the control-like cohort, finding that prioritising variants with at least one pathogenic/likely pathogenic classification and no more than one benign/likely benign classification yielded a specificity of 97% with minimal impact on sensitivity (Supplemental Figure 2). Variants prioritised in multiple individuals were manually curated, identifying low penetrance variants and recurrent artefacts, which were subsequently added to the exclusion list, increasing specificity by ∼5%.

### Genes included with multiple modes of inheritance

Genes included under multiple modes of inheritance led to a disproportionate number of non-reportable variants as mode of inheritance filters are applied at a gene level and not a variant level. Initially the three genes with the lowest specificity (*ALPL, CHRNE, ABCC8*) were all included under autosomal dominant and recessive inheritance (Supplemental Table 10). This led to many individuals with prioritised heterozygous variants that are only pathogenic if recessive and a reduction of overall specificity by ∼1.6%. These genes are now included in the study under only autosomal recessive mode of inheritance.

### Presence of specific variants only clinically relevant in combination

Variants clinically relevant only as part of a complex allele may be prioritised without all component alleles. The *BTD* variant NM_001370658.1:c1270G>C was prioritised in 123 samples in the control-like cohort (Supplemental Table 11), including 64 in a homozygous state^24^, however this variant is not considered pathogenic unless compound heterozygous with another variant. The variant was prioritised in 39 samples as potentially compound heterozygous with NM_001370658.1:c.451G>A. This has been observed in affected individuals when found in *cis* and in *trans* with a third variant^25,26^. Similarly, 20 samples were prioritised with NM_000400.4:c.2150C>G and NM_000400.4:c.1381C>G in *ERCC2*, typically reported in *cis*^27–31^ and requiring a third variant in *trans* to be considered pathogenic^29^. Clinical scientists reviewing these cases will be provided guidance on frequent variant combinations. The *BTD* variant NM_001370658.1:c.1270G>C will be added to the exclusion list as it is not reportable in isolation but if NM_001370658.1:c.451G>A is prioritised then clinical scientists will be able to check the genotype manually.

### Variants prioritised in overlapping and nearby genes

Variant matching for inclusion list variants is done by genomic coordinates and alleles which may result in inappropriate prioritisation of variants in overlapping or nearby genes. While few overlapping gene variants were prioritised in our cohort, all potential overlapping ClinVar variants were added to the exclusion list.

### Targeted callers for genes in challenging regions of the genomes

Targeted callers in DRAGEN v4 were introduced for genes in complex genomic regions, including *SMN1/2* and *CYP21A2*. These were individually assessed to ensure high specificity. The *SMN* copy number caller was tested on 75,582 rare disease participants from the 100,000 Genomes Project. Eight participants were identified with homozygous *SMN1* loss, three of whom had a confirmed *SMN1* deletion, while the remaining five had muscular phenotypes, suggesting no false positives.

The *CYP21A2* caller was tested on the control-like subset using an internal inclusion list to prioritise only relevant variants. Six unique variants were prioritised across 12 samples (∼0.2% of the cohort), a >30 fold enrichment of what we would expect given the estimated prevalence of congenital adrenal hyperplasia in the UK^32^. Given the complexity of the region, these variants are not able to be manually assessed through the Integrative Genomics Viewer^33^, therefore the decision was made to remove the gene from the study to avoid false positives. Improvements to the caller may allow this gene to be included in the future.

#### Estimating the specificity of variant prioritisation

After addressing these issues, specificity across the control-like cohort was estimated at 96.3% (95% Wilson CI: 96.05%, 96.45%) (Table 1). This cohort was aligned using a previous version of DRAGEN than will be used in the study and so specificity was also calculated on a subset of 5,855 samples realigned with DRAGEN v4 yielding a similar estimate of 97.3% (95% CI: 96.78%,97.62%). Specificity was comparable across prioritisation sources and all genes maintained a specificity >99.95% (Supplemental Figure 3a,c).

There were 334 samples with potential compound heterozygous variants prioritised in the control-like cohort. We queried the co-occurrence of these variants in gnomAD and found that for the 242 samples we were able to query, 69% of prioritised variants were predicted to be on the same haplotype, 26% on different haplotypes and 5% were uncertain (Supplemental Table 12). Manual curation of all prioritised variants in the control-like subset estimated that 35 samples would have a reportable variant, 0.6% of the cohort. Seven of these samples had unphased potential compound heterozygous variants. Likelihood of the co-occurrence of these variants was queryable in gnomAD for four samples which were all predicted to be on different haplotypes. The conditions associated with the reportable variants in this cohort were plausible, as although the cohort was not enriched for rare diseases, individuals with such conditions may still be present (Supplemental Table 13).

Specificity was further estimated in a separate cohort of 1,362 Cancer GMS samples, yielding a similar result of 95.5% (95% Wilson’s CI: 94.21%-96.43%). Comparing gene specificity between the control-like and replication cohorts revealed a significant enrichment of prioritised variants in cancer-related genes (12/1,362 replication cohort vs. 117/34,410 control-like cohort, proportion test p=0.00109), which may explain the reduced specificity. No significant difference was found for non-cancer-related genes (50/1,362 replication cohort vs. 1,178/34,410 control-like cohort, proportion test p=0.677) (Supplemental Figure 3b).

#### Sensitivity of variant prioritisation in individuals with rare genetic disease

We analysed 545 rare disease participants from the NHS GMS and 100,000 Genomes Project with diagnostic pathogenic/likely pathogenic small variants in newborn genes aligned with the mode of inheritance included in the study. This dataset consisted of 573 unique variants across 172 different genes. There were genes which contributed a much larger number of variants than others. In the 100,000 Genomes Project participants there were many *COL1A1/2* variants likely due to recruitment strategies (Figure 3). These samples were then run through variant prioritisation. Prioritisation through presence in CVA was ignored since this was the source of variants. We found that 80.8% of samples had at least one diagnostic variant prioritised. The source of prioritisation was well distributed (Supplemental Figure 4) and sensitivity estimates were similar in monoallelic and biallelic genes (Supplemental Table 13).

**Figure 3:**
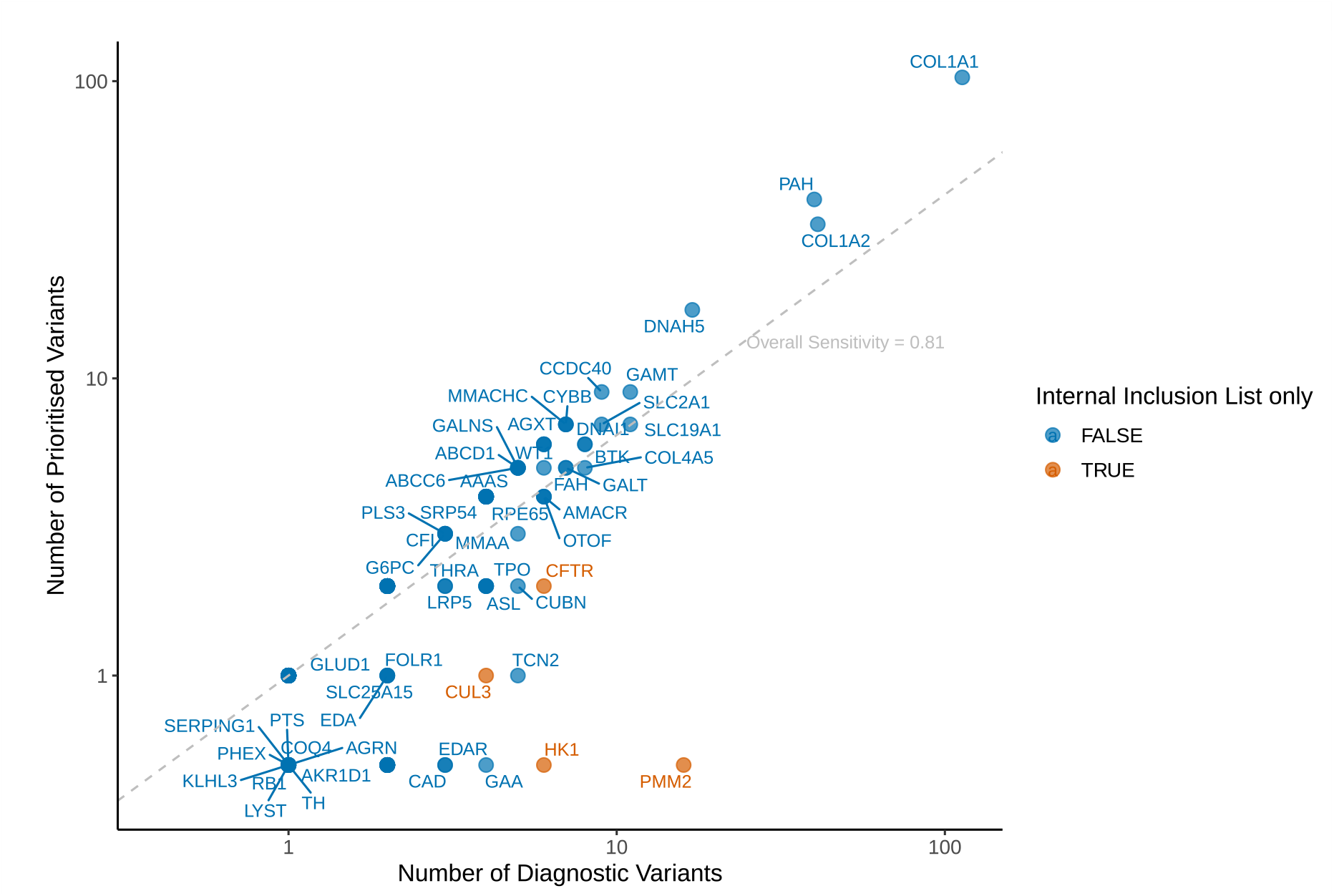
Sensitivity of automated variant prioritisation in the Generation Study across diagnostic variants from participants in the 100,000 Genomes Project and NHS Genomic Medicine Service. Number of diagnostic variants per gene against how many of these would be prioritised for clinical scientist review. Coloured by whether the gene will only have variants prioritised through an internal inclusion variant list.

Unprioritised diagnostic variants were mostly missense variants absent from ClinVar or QIAGEN (Supplemental Table 14). Diagnostic variants in genes where only internal inclusion list variants are prioritised were less frequently prioritised, resulting in lower sensitivity estimates (Supplemental Table 15, Figure 3a). Reviewing reported phenotypes, 24/25 participants with diagnostic variants in internal inclusion list-only genes did not have newborn-relevant phenotypes. Eighteen out of these 24 participants with diagnostic variants in *PMM2* where their phenotypes aligned with congenital disorder of glycosylation, type Ia rather than Polycystic kidney disease with hyperinsulinemic hypoglycemia which is included in the Generation Study. There was a single participant with cystic fibrosis where a diagnostic *CFTR* variant was not prioritised, however it was reported that this individual additionally had a LINE insertion in the gene which we would not currently be able to prioritise in the pipeline.

## Discussion

Genomic sequencing offers the opportunity to screen for hundreds of rare conditions with a single test. In newborn screening on a population level, high specificity is essential to minimise unnecessary investigation and intervention and reduce the burden on clinical services whilst maintaining sufficient sensitivity. This is especially important at the beginning stages of the study as initial results inform refinements. Our automated variant prioritisation approach streamlines screening, reducing the clinical scientist workload and ensuring a quick turnaround, critical when scaling to a population level.

We estimate that 3-5% of individuals will have a prioritised variant requiring manual review, with <1% having a reportable variant. These individuals would undergo non-genomic confirmatory testing and genetic confirmation where possible. The estimated proportion of reportable variants is lower than initial estimates from similar studies such as the GUARDIAN study^12^. However, these differences are largely attributable to the genes included in the respective studies, such as the absence of *G6PD* in the Generation Study, rather than substantial differences in the analysis approach. Among participants with rare genetic disease, variant prioritisation identified ∼80% of diagnostic variants in genes included in the Generation Study.

There are limitations to these estimates. For participants with diagnostic variants in newborn genes, we did not confirm whether these cases have phenotypes that are relevant to newborns and could bias this estimate downwards. Conversely the fact that these variants already deemed diagnostic are more likely to be present in ClinVar and prioritised. Moreover, datasets like QIAGEN, which are less widely used in variant classification, may be undervalued. Selection bias may also be present, with certain genes disproportionately represented while others are absent. Additionally, spectrum bias may arise as more straightforward cases were less likely to undergo GS in the NHS Genomic Medicine Service. Although specificity estimates are derived from a cohort not expected to be enriched for rare diseases, the presence of such conditions cannot be excluded, as evidenced by the enrichment of prioritised variants in cancer-associated genes in the replication cohort. While the specificity cohort reflects the composition of the British population, we lack a good understanding of how these estimates may differ across individuals of non-European genetic ancestry, particularly those underrepresented in genomic studies. Similarly, the sensitivity cohort may be subject to ancestry biases and may not adequately represent all ancestries in Britain. To understand this impact, variant prioritisation approaches should be validated in genetically diverse cohorts. Addressing these limitations will require greater representation of diverse ancestries in reference databases such as gnomAD and ClinVar as well as enhancing these resources with population-specific haplotype data.

Improving variant prioritisation will be an iterative process. We will closely monitor outputs and can remove genes/variants accordingly. The challenges encountered highlight areas for improvement. The implementation of variant specific rules may aid prioritisation of complex alleles. Adjusting allele frequency thresholds for different modes of inheritance beyond pLoF prioritisation may allow the reintroduction of conditions where genes are considered under multiple modes of inheritance. Future iterations will incorporate other types of variation such as structural variants and STRs. Population co-occurrence data may improve identification of compound heterozygous variants, but further validation is required to assess performance across genetic ancestries before implementation.

Overall, we have provided a framework for variant prioritisation in newborn screening and demonstrated its potential to identify disease-causing variants whilst maintaining high specificity. Our results highlight the importance of evaluating analytical validity for each gene before inclusion in screening. Understanding the proportion of samples with prioritised variants is crucial for service design, informing workforce requirements for variant review and follow-up. As results accumulate from the Generation Study, they will further clarify the true positive predictive value and clinical utility of this approach.

## Data Availability

Data from the National Genomic Research Library (NGRL) supporting this study are available within the secure Genomics England Research Environment. Data used or referred to in this publication include: 100,000 Genomes Project data from Rare Disease arm, 100,000 Genomes Project data from Cancer arm, Covid-19 aggregate data, NHS Genomic Medicine Service data for rare disease participants. Access is restricted to approved researchers who are members of the Genomics England Research Network, subject to a data access agreement and participant-led governance. For more information on data access, visit: https://www.genomicsengland.co.uk/research

## Ethics Declaration

The Generation study has been approved by the Health Research Authority and Cambridge Central Research Ethics Committee. Clearance number: 324562.

The NGRL is approved by the East of England – Cambridge Central Research Ethics Committee (REC reference: 20/EE/0035)

Informed consent was obtained from all participants.

## Declaration of AI and AI-assisted technologies in the writing process

During the preparation of this work the author(s) used ChatGPT to assist with editing for clarity and conciseness. After using this tool/service, the author(s) reviewed and edited the content as needed and take(s) full responsibility for the content of the publication

## Acknowledgements

The Generation Study is funded by the UK Department of Health and Social Care. This research was made possible through access to data in the National Genomic Research Library, which is managed by Genomics England Limited (a wholly owned company of the Department of Health and Social Care). The National Genomic Research Library holds data provided by patients and collected by the NHS as part of their care and data collected as part of their participation in research. The National Genomic Research Library is funded by the National Institute for Health Research and NHS England. The Wellcome Trust, Cancer Research UK and the Medical Research Council have also funded research infrastructure.

## Author contributions

Conceptualization: [D.K., A.T.L., E.T., R.S.];

Data curation: [G.A., M.A.G., A.Pu., I.L.,]

Formal analysis: [J.K., P.S., D.D, K.Sa.];

Funding acquisition: [A.T.L., E.T., R.S.];

Investigation: [M.A.G., A.Pu., I.L., D.B, K.St., M.T., A.Pi];

Methodology: [D.K., J.K., P.S., D.D., E.B., A.Pa];

Software: [J.S., P.Q., J.K., P.S., D.D];

Supervision: [D.K.]; Visualisation: [J.K.];

Writing – original draft: [J.K.];

Writing – review & editing: [J.K., D.K., D.D., P.S., A.Pu., K.Sa, E.B., M.T., K.St., D.B., A.Pi., E.T.]

