## Supplemental Tables/Figures for "Assessment of the variant prioritisation strategy for genomic newborn screening in the Generation Study"

### Supplemental Figures and Tables

### Figures

**Supplemental Figure 1:** Coverage metrics per gene across the control-like subset aligned to DRAGEN v4 and across all exons.. Genes with metrics below the shown thresholds are labeled. For the figures b,d and f which show metrics for all exons, the corresponding gene is labelled as opposed to the exact exon for readability. The metrics are as follows: a) median coverage per gene (b) median coverage per exon (c) mean coverage per gene (d) mean coverage per exon (e) proportion of gene with coverage >15X (f) proportion of exon with coverage >15X .

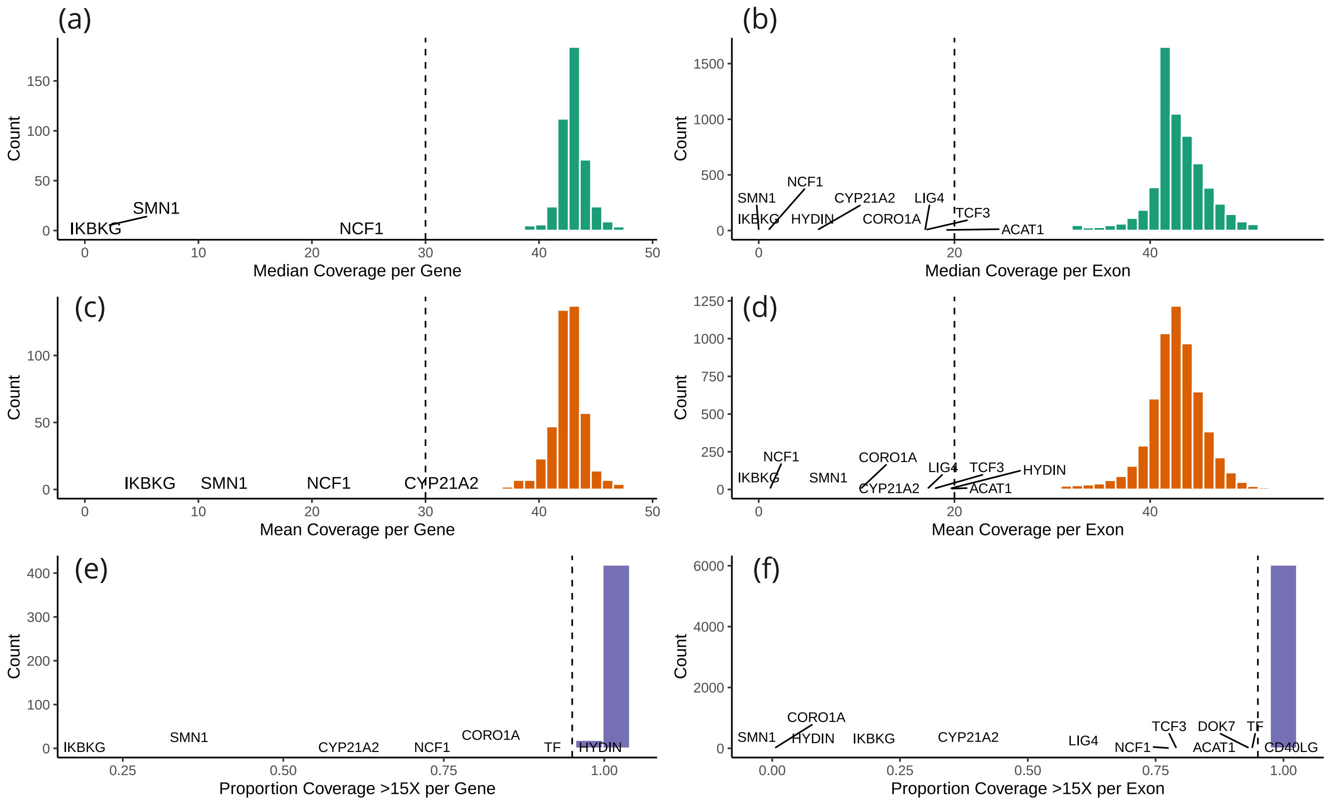

**Supplemental Figure 2:** Effect of different prioritisation rules for ClinVar variants with conflicting interpretations of pathogenicity (CIP). (a) Specificity of automated variant prioritisation in the control-like cohort (b) Sensitivity of automated variant prioritisation in the participants with diagnostic variants in genes included in the Generation Study from 100,000 Genomes Project and NGS Genome Medicine Service. Rules applied are detailed in the methods, thresholds on number of classifications. (P/LP = Pathogenic/Likely Pathogenic, B/LB = Benign Likely Benign)

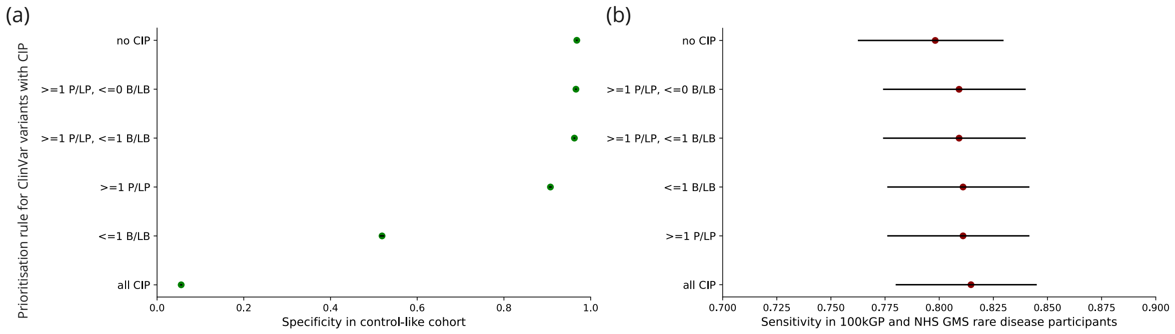


**Supplemental Figure 3:** Specificity of automated variant prioritisation. Comparing gene specificity in control-like cohort (DRAGEN v3.2) and (a) the control like subset realigned with DRAGEN v4.0 (b) replication cohort which is made up of participants recruited with cancer. The colour indicates whether the gene is also associated with cancer. (c) The number of samples with variants prioritised in the control-like cohort by prioritisation source.

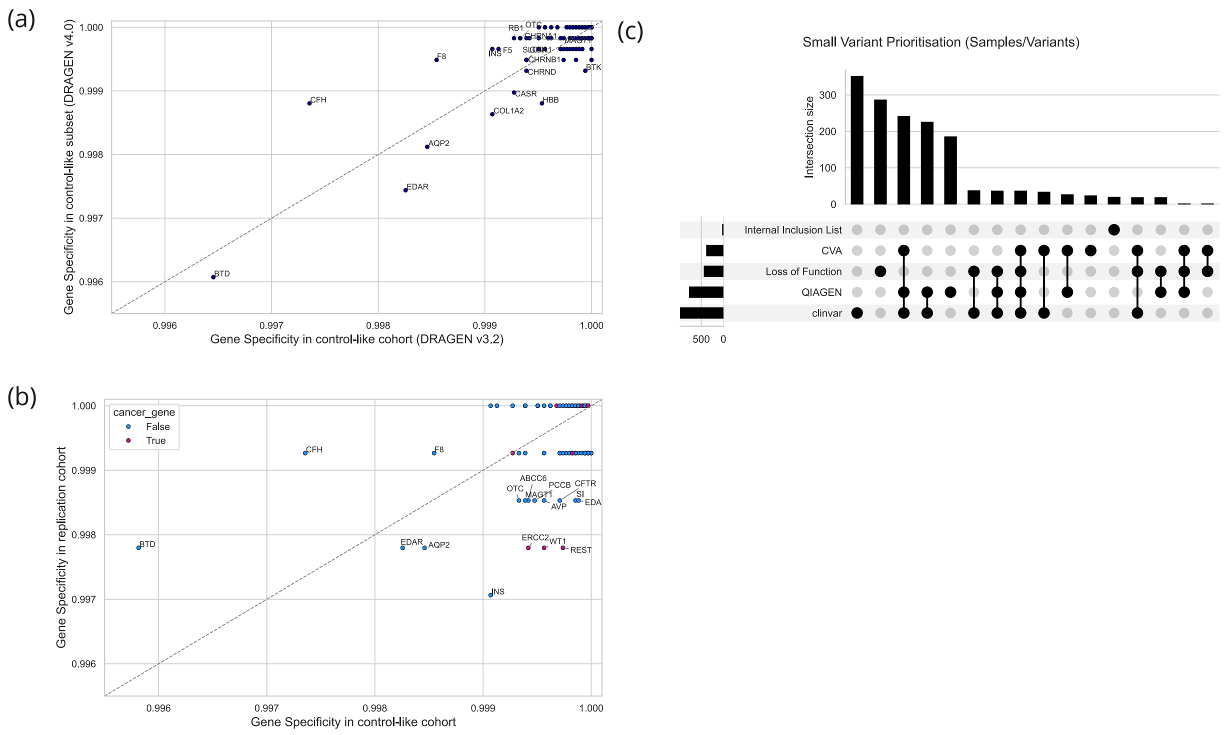

**Supplemental Figure 4:** Counts of prioritised diagnostic variants by prioritisation source in the 100,000 Genome Project and NHS GMS rare disease participants.

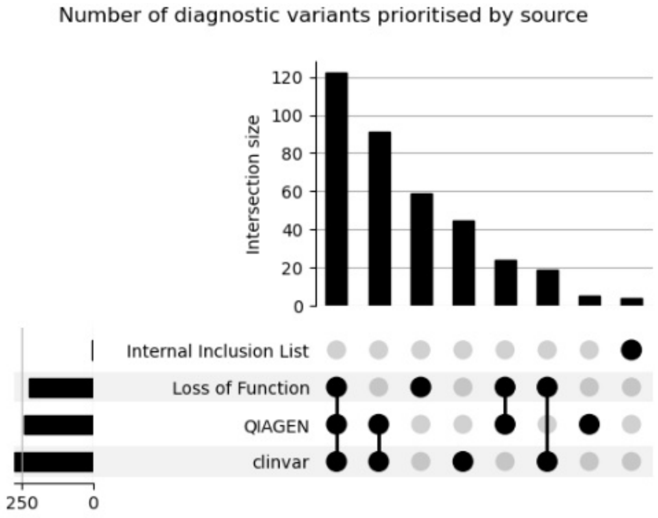

Tables

**Supplemental Table 1:** Principles for selecting conditions in the Generation Study

| Principle | Description | Additional considerations |
|---|---|---|
| A | There is strong evidence that the genetic variant(s) causes the condition and can be reliably detected. | Where appropriate, there may be a confirmatory test that can establish whether or not the child has the condition. |
| B | A high proportion of individuals who have the genetic variant(s) would be expected to have symptoms that would have a debilitating impact on quality of life if left undiagnosed. | The impact on quality of life should consider factors such as the testimony of patients and families affected including social and environmental factors, and quality-adjusted life years (QALYs) where available. |
| C | Early or pre-symptomatic intervention for the condition has been shown to lead to substantially improved outcomes in children, compared to intervention after the onset of symptoms. | The intervention would normally be initiated in early childhood (by age 5); and could either cure, delay or modify the course of the condition. |
| D | Conditions screened for are only those for which the interventions are equitably accessible for all. | Incorporating input from NHS England and other relevant clinical and commissioning bodies. |

**Supplemental Table 2:** Gene-condition pairs assessed for analytical validity (CSV file)
gene_name: HGNC gene name.
newborn_phenotype: Associated phenotype considered for inclusion in the Generation Study.
mode_of_inheritance: Mode of inheritance for the associated condition.
location_GrCH38: Genomic location of the gene (GRCh38).
ensembl_id : Ensembl gene ID.
included_in_GS: whether the gene-condition pair was included in the Generation Study.
internal_inclusion: whether there is an internal inclusion variant list only for this gene
lof_prioritisation: whether loss-of-function variant prioritisation was implemented in the pipeline.
targeted_caller: whether a targeted variant caller was assessed for this gene.

**Supplemental Table 3:** Ancestry composition of control-like cohort compared to the UK Office of National Statistics (ONS) live births occurring in 2020 by ethnicity (https://www.ons.gov.uk/). In the ONS statistics: European include White British and White Other, South Asian includes Bangladeshi, Indian and Pakistani, African includes Black African and Black Carribean and any other Black background. The ancestry projections were done by projecting principal components onto 1000 Genomes data (European (EUR), South Asian (SAS) and African (AFR)).

| Ancestry | Control-like cohort inferred ancestry | ONS 2020 Live births (reported) |
|---|---|---|
| European | 78.2% | 70.6% |
| South Asian | 7.8% | 9.9% |
| African | 4.3% | 5.1 % |

**Supplemental Table 4:** Coverage and CNV callability metrics for all genes considered (tsv file). Below is a description of each column:
hgnc_symbol: HGNC-approved gene symbol.
ensembl_gene_id: Unique Ensembl Gene ID for the gene.
ensembl_transcript_id: Unique Ensembl Transcript ID specifying the assessed transcript.
location: Genomic coordinates (chromosome and start/end positions) of the gene or transcript.
total_bases: Total number of bases assessed within the transcript.
total_exons: Total number of exons in the transcript.
bases_less_10: Number of bases in the transcript with coverage below 10X.
percentage_less_10: Percentage of total bases in the transcript with coverage below 10X.
num_exons_less_10: Number of exons in the transcript where median coverage falls below 10X.
bases_less_15: Number of bases in the transcript with coverage below 15X.
percentage_less_15: Percentage of total bases in the transcript with coverage below 15X.
num_exons_less_15: Number of exons in the transcript where median coverage is below 15X.
bases_less_20: Number of bases in the transcript with coverage below 20X.
percentage_less_20: Percentage of total bases in the transcript with coverage below 20X.
num_exons_less_20: Number of exons in the transcript where median coverage is below 20X.
bases_less_25: Number of bases in the transcript with coverage below 25X.
percentage_less_25: Percentage of total bases in the transcript with coverage below 25X.
num_exons_less_25: Number of exons in the transcript where median coverage is below 25X

**Supplemental Table 5:** Internal Variant Inclusion List (csv file)
Columns: Chromosome, Position, Reference, Alternate, Published variant nomenclature, HGVS, Evidence (PMID to support inclusion of variant), Gene (HGNC)

**Supplemental Table 6:** Variant exclusion list (csv file)
Columns: Gene (HGNC), Chromosome, Position, Reference, Alternate, HGVS

**Supplemental Table 7:** Gene-condition pairs with internal inclusion lists included in the Generation Study. pLoF prioritisation enabled describes whether pLoF variants are prioritised in this gene as well. Table describes gene specificity using the regular rules (without internal inclusion list) and with internal inclusion list.

| Gene | Condition | Mode of Inheritance | Mechanism of Pathogenicitiy | pLoF prioritisation enabled | Reason for internal inclusion list | Gene Specificity without internal inclusion list (%) | Gene Specificity with internal inclusion list (%) |
|---|---|---|---|---|---|---|---|
| *ALAS2* | Protoporphyria, erythropoietic, X-linked | x-linked biallelic | gain of function | No | Do not want to prioritise variants in this gene that are associated with Anemia, sideroblastic, 1, | 99.93 | 100 |
| *CFTR* | Cystic fibrosis | biallelic | loss of function | Yes | Do not want to prioritise variants in this gene that are associated with other CFTR-related disorders such as Congenital Bilateral Absence of the Vas Deferens. | 97.81 | 99.97 |
| *CUL3* | Pseudohypoaldosteronism, type IIE | monoallelic | dominant negative | No | Do not want to prioritise variants in this gene that are associated with a neurodevelopmental disorder that is not included which is associated with LoF. | 100 | 100 |
| *HK1* | HK1 associated hyperinsulinism | monoallelic | gain of function | No | Do not want to prioritise variants in this gene that are associated with other conditions we are not including. | 99.71 | 100 |
| *IFITM5* | Osteogenesis Imperfecta type V | monoallelic | gain of function | No | Only 1 specific variant (c.-14C>T) has been associated with OI | 100 | 100 |

| | | | | | | | |
|---|---|---|---|---|---|---|---|
| | | | | | type V. Another missense variant (S40L), for e.g. has been associated with type VI, which we do not want to prioritise. | | |
| *KCNJ11* | Diabetes, permanent neonatal 2, with or without neurologic features Familial hyperinsulinemic hypoglycemia-2 | monoallelic biallelic | gain of function loss of function | Yes | Do not want to prioritise variants in this gene that are associated with other conditions. The internal inclusion list is specifically for monoallelic "Diabetes, permanent neonatal 2, with or without neurologic features" as the mechanism of pathogenicity is gain of function (GoF). LoF algorithm will still be enabled for KCNJ11 for biallelic "Familial hyperinsulinemic hypoglycemia-2". | 99.93 | 100 |
| *NLRP3* | Cryopyrin associated periodic fever syndrome | monoallelic | gain of function | No | Do not want to prioritise variants associated with other conditions in this gene | 99.99 | 99.99 |
| *PMM2* | Polycystic kidney disease with hyperinsulinemic hypoglycemia | biallelic | gain of function | No | Do not want to prioritise variants in this gene that are associated with Congenital disorder of glycosylation type 1a | 99.99 | 100 |

| | | | | | | | |
|---|---|---|---|---|---|---|---|
| *RAC2* | Immunodeficiency 73B | monoallelic | gain of function | No | Don't want to prioritise variants in this gene associated with Immunodeficiency 73A and Immunodeficiency 73C | 100 | 100 |
| *SAMD9* | MIRAGE syndrome | monoallelic | gain of function | No | Do not want to prioritise variants in this gene that are associated with other conditions such as adult-onset myelodysplastic syndrome. | 99.97 | 99.99 |
| *WNK1* | Pseudohypoaldosteronism, type IIC | monoallelic | gain of function | No | Do not want to prioritise variants in this gene that are associated with neuropathy | 99.92 | 100 |
| *CDKN1C* | IMAGE syndrome | monoallelic | gain of function | No | Do not want to prioritise variants in this gene that are associated with Beckwith-Wiedemann syndrome | 100 | 100 |

**Supplemental Table 8:** Allele frequency thresholds for gnomAD populations in pLoF prioritisation (gnomAD Genomes v3.1.2, gnomAD Exomes v2.1.1). These differ for gene-conditions associated with dominant vs recessive phenotypes.

| Dataset | Population | Dataset size (individuals) | Monoallelic (dominant) | Biallelic (recessive) |
|---|---|---|---|---|
| Internal | Mixed | 5,855 | 0.001 | 0.01 |
| GNOMAD_GENOMES | African/African American | 20,744 | 0.0005 | 0.01 |
| GNOMAD_GENOMES | Latino/Admixed American | 7,647 | 0.001 | 0.01 |
| GNOMAD_GENOMES | Ashkenazi Jewish | 1,736 | 0.003 | 0.01 |
| GNOMAD_GENOMES | East Asian | 2,604 | 0.002 | 0.01 |
| GNOMAD_GENOMES | European (Finnish) | 5,316 | 0.001 | 0.01 |
| GNOMAD_GENOMES | Middle Eastern | 158 | 0.1 | 0.1 |
| GNOMAD_GENOMES | European (non-Finnish) | 34,029 | 0.0005 | 0.01 |
| GNOMAD_GENOMES | South Asian | 2,419 | 0.002 | 0.01 |
| GNOMAD_EXOMES | Latino/Admixed American | 17,296 | 0.0005 | 0.01 |
| GNOMAD_EXOMES | Ashkenazi Jewish | 5,040 | 0.001 | 0.01 |
| GNOMAD_EXOMES | East Asian | 9,197 | 0.001 | 0.01 |
| GNOMAD_EXOMES | European (Finnish) | 10,824 | 0.001 | 0.01 |
| GNOMAD_EXOMES | European (non-Finnish) | 56,885 | 0.0005 | 0.01 |
| GNOMAD_EXOMES | South Asian | 15,308 | 0.001 | 0.01 |

**Supplemental Table 9**: Gene specificity across genes included in the Generation Study in the control-like cohort ('gene_specificity_controllike'), control-like subset ('gene_specificity_controllike_subset') and the replication cohort ('gene_specificity_replication'). Note that this only contains genes where a variant is prioritised, all genes not included in this table have specificity of 1 across all cohorts.

**Supplemental Table 10:** Genes and conditions included under different modes of inheritance and impact on gene specificity. Table describes genes where the modes of inheritance included in the Generation Study were changed due to issues in gene specificity.

| Gene | Conditions included | Modes of Inheritance included | Number of prioritised variants | Number of prioritised samples | Gene specificity (%) |
|---|---|---|---|---|---|
| *CHRNE* | Myasthenic syndrome, congenital, 4, autosomal recessive; Myasthenic syndrome, congenital, 4, autosomal dominant | monoallelic and biallelic | 25 | 219 | 99.37 |
| | Myasthenic syndrome, congenital, 4, autosomal recessive | biallelic | 2 | 1 | 99.99 |
| *ALPL* | Autosomal recessive hypophosphatasia; Autosomal dominant hypophosphatasia | monoallelic and biallelic | 55 | 183 | 99.47 |
| | Autosomal recessive hypophosphatasia | biallelic | 5 | 3 | 99.99 |
| *ABCC8* | Generalized arterial calcification of infancy 2; Diabetes mellitus, permanent neonatal 3, autosomal recessive; Hyperinsulinemic hypoglycemia, familial, 1; Diabetes mellitus, permanent neonatal 3, autosomal dominant; ABCC9 associated hypertrichotic osteochondrodysplasia | monoallelic and biallelic | 71 | 172 | 99.50 |
| | Generalized arterial calcification of infancy 2 ; Diabetes mellitus, permanent neonatal 3, autosomal recessive; Hyperinsulinemic hypoglycemia, familial, 1 | biallelic | 0 | 0 | 100 |

**Supplemental Table 11:** Variants prioritised in the largest number of samples in the control-like cohort

| Variant ID (chrom_pos_ref_alt) | HGVS | Gene | Prioritisation Source | Number of samples with variant prioritised |
|---|---|---|---|---|
| 3_15645186_G_C | NM_001370658.1:c.1270G>C | *BTD* | CVA;clinvar;QIAGEN | 123 |
| 1_196749030_196832721 | NC_000001.11:g.196749030_196832721del | *CFH* | Loss of Function | 62 |
| 3_15644367_G_A | NM_001370658.1:c.451G>A | *BTD* | CVA;clinvar;QIAGEN | 39 |
| 1_169549811_C_T | NM_000130.5:c.1601G>A | *F5* | clinvar | 30 |
| 2_108930972_C_T | NM_022336.4:c.43G>A | *EDAR* | clinvar | 30 |
| 19_45357368_G_C | NM_000400.4:c.1381C>G | *ERCC2* | CVA;QIAGEN | 20 |
| 3_136327178_AT_A | NM_000532.5:c.1223del | *PCCB* | Loss of Function;clinvar | 18 |
| 19_45352249_G_C | NM_000400.4:c.2150C>G | *ERCC2* | CVA;clinvar;QIAGEN | 18 |
| 3_136327173_G_GTA | NM_000532.5:c.1217_1218insTA | *PCCB* | Loss of Function | 18 |
| 3_136327181_ATCC_A | NM_000532.5:c.1226_1228del | *PCCB* | CVA | 18 |

**Supplemental Table 12:** Potential compound heterozygous variants prioritised in the control-like cohort broken down by co-occurrence prediction in gnomAD. This is also subset to those that are >150bp apart.

| Co-occurrence prediction | All potential compound heterozygous variants | | Potential compound heterozygous variants >150bp apart | |
|---|---|---|---|---|
| | Number of samples | Number of unique variant pairs | Number of samples | Number of unique variant pairs |
| Different haplotye | 64 | 45 | 50 | 40 |
| Same haplotype | 167 | 36 | 109 | 26 |
| Uncertain | 11 | 7 | 6 | 6 |
| No prediction | 100 | 69 | 49 | 39 |

**Supplemental Table 13:** Conditions associated with variants deemed reportable in control-like subset.

| Gene | Zygosity of reportable variant | Condition |
|---|---|---|
| *ABCD1* | homozygous | Adrenoleukodystrophy |
| *AVP* | heterozygous | Diabetes insipidus, neurohypophyseal |
| *BTK* | homozygous | X-linked Agammaglobulinaemia |
| *CCDC103* | homozygous | Ciliary dyskinesia, primary, 17 |
| *COL1A1* | heterozygous | COL1A1 related Osteogenesis Imperfecta |
| *COL1A2* | heterozygous | COL1A2 related Osteogenesis Imperfecta |
| *DUOX2* | homozygous | Thyroid dyshormonogenesis 6 |
| *EDA* | homozygous | Ectodermal dysplasia 1, hypohidrotic, X-linked |
| *EDAR* | heterozygous | Ectodermal dysplasia 10A, hypohidrotic/hair/nail type, autosomal dominant |
| *EDARADD* | heterozygous | Ectodermal dysplasia 11B, hypohidrotic/hair/tooth type, autosomal recessive |
| *F7* | homozygous | Factor VII deficiency |
| *F8* | homozygous | Haemophilia A |
| *HBB* | homozygous | Sickle Cell Disease |
| *IRS4* | homozygous | Hypothyroidism, congenital, nongoitrous, 9 |
| *KLHL3* | heterozygous | Pseudohypoaldosteronism, type IID, autosomal dominant |
| *PAH* | homozygous | Phenylketonuria |
| *PAX8* | heterozygous | Hypothyroidism, congenital, due to thyroid dysgenesis or hypoplasia |
| *RAPSN* | homozygous | Congenital myasthenic syndrome-11 |
| *RPL11* | heterozygous | Diamond-Blackfan anaemia 7 |
| *RPS26* | heterozygous | Diamond-Blackfan anaemia 10 |
| *SLC2A1* | heterozygous | GLUT1 deficiency syndrome-1, Autosomal Dominant |
| *THRB* | heterozygous | Thyroid hormone resistance, autosomal dominant |
| *AGPAT2* | compound heterozygous | Congenital generalized lipodystrophy type 1 |
| *AGXT* | compound heterozygous | Hyperoxaluria, primary, type 1 |
| *CBS* | compound heterozygous | Homocystinuria, B6-responsive and nonresponsive types |
| *CFTR* | compound heterozygous | Cystic fibrosis |
| *CTNS* | compound heterozygous | Cystinosis, nephropathic |

| | | |
|---|---|---|
| *CYP27A1* | compound heterozygous | Cerebrotendinous xanthomatosis |
| *HBB* | compound heterozygous | Sickle Cell Disease |
| *PAH* | compound heterozygous | Phenylketonuria |

**Supplemental Table 14:** Sensitivity by mode of inheritance for 100kGP and NHS GMS rare disease participants with diagnostic variants in genes included in the Generation Study. Sensitivity also split by genes with internal inclusion lists.

| Mode of Inheritance for condition included | Internal Inclusion list only genes | Total number of diagnostic variants | Number of prioritised diagnostic variants | Total samples | Number of Samples with >=1 variant prioritised | Sensitivity (%) |
|---|---|---|---|---|---|---|
| biallelic | False | 411 | 326 | 279 | 230 | 82.44 |
| | True | 22 | 2 | 12 | 2 | 16.67 |
| monoallelic | False | 194 | 169 | 194 | 169 | 87.11 |
| | True | 13 | 2 | 13 | 2 | 15.38 |
| X-linked biallelic | False | 41 | 33 | 41 | 33 | 80.49 |
| X-linked monoallelic | False | 6 | 5 | 6 | 5 | 83.33 |

**Supplemental Table 15:** Predicted consequences of diagnostic variants that were not prioritised in 100kGP and NHS GMS rare disease participants with diagnostic variants in genes included in the Generation Study

| Predicted Consequence of diagnostic variant | Number of variants not prioritised |
|---|---|
| Missense | 54 |
| Splice region | 8 |
| Inframe deletion | 2 |
